# Discerning knowledge and practices towards diabetic foot care among adult diabetic patients attending St. Mary’s Mission Hospital, Nairobi. Insight into managing a potential diabetic foot

**DOI:** 10.1101/2025.08.27.25334604

**Authors:** Norah Anne Moggute Oyagi, Okubastion Tekeste Okube, Magdalene Philip Umoh, Abraham Isiaka Jimmy, Lee Presley Gary

**Author notes:** Corresponding Author (Lee Presley Gary, Jr.). Holy Spirit Hospital - Masuba, Makeni, Northen Province, Sierra Leone (Norah Anne Moggute Oyagi). College of Health Sciences, University of Moi, Eldoret, Kenya (Abraham Isiaka Jimmy). Strategic Management Services – USA, New Orleans. Louisiana, USA. EMAIL ADDRESSES: Norah Anne Mogute Oyagi) Okubastion Tekeste Okube Magdalene Philip Umoh Abraham Isiaka Jimmy. CONSENT for PUBLICATION: All authors have acknowledged, agreed, and joined *in toto* for the publication of this research project and submission of the corresponding manuscript to MED RX IV.

## Abstract

In our research study, we investigated how well people living with diabetes mellitus optimize the overall management of their condition to prevent development of diabetic foot or feet. Such vital limb complications, arising from poor knowledge and practices on foot care, especially in low socioeconomic households, are some of the serious outcomes of poorly managed diabetes mellitus.

Our study sought out and identified the favorable factors that can help mitigate adult diabetic foot.

By design, we surveyed and assessed the level of knowledge and practices towards diabetic foot care among adult diabetic patients attending a Medical Outpatient Clinic at a faith-based (Level IV) hospital in Nairobi, Keyna. A descriptive, cross-sectional hospital-based study was detailed with authorized permits and launched in May 2021, involving 140 adults living with diabetes who attended the cooperating institution, St. Mary’s Mission Hospital in Nairobi. The study participants were recruited by a census method. Data was collected on the level of knowledge and practices towards diabetic foot care using a pretested questionnaire for adult diabetic patients. Data analysis was done using descriptive and inferential statistics. Out of the 140 participants, 66% demonstrated a low level of knowledge on diabetic foot care and only 8% had a high level of knowledge. Half of the participants did not practice foot care as medically recommended. We found no significant association between the practice of adult diabetic foot care and socio-demographic variables. The findings reveal that both the level of knowledge and practice towards foot care by adults attending St. Mary’s Mission Hospital with diabetic mellitus conditions was strikingly unsatisfactory and served as a precursor for poor health.

## 1. INTRODUCTION

Diabetes mellitus(DM) is a chronic condition that poses significant risks to lower limb health, particularly through the development of diabetic foot ulcers (DFUs). Diabetic foot is because of complicated or poorly managed diabetes mellitus. It could lead to a chronic wound or even loss of limb(s) in surgical amputations and an immense psychological, socio-economic burden not only to the individual patient, but also to their families and even public health systems [1]. These complications often arise due to peripheral neuropathy, poor circulation, and delayed wound healing, which can lead to infection, gangrene, and even amputation if not properly managed [2].

A recently published report indicated that up to 85% of diabetic foot complications are preventable through proper education, regular foot inspections, and timely medical intervention [3]. Despite the availability of solid evidence and clinical guidelines, many diabetic patients lack adequate knowledge and fail to practice proper foot care. Research conducted in various regions, including Kenya, Saudi Arabia, and Iran, reveals that while some patients are aware of diabetic foot risks, many do not translate this knowledge into consistent foot care practices [3,4,5].

In Kenya, a study found that adult diabetic patients often had limited understanding of foot care and engaged in risky behaviors such as walking barefoot or neglecting foot inspections. Similar findings were reported in Embu County, Kenya, where poor foot care practices were significantly associated with foot ulcer development [5].

This gap in awareness and behavior contributes significantly to the burden of diabetic foot complications, especially in low-resource settings. Research has shown that poor foot care knowledge and practices are strongly linked to increased risk of diabetic foot ulcers and subsequent complications [3]. Understanding patient behavior and knowledge can guide targeted education programs and clinical interventions.

The main objective of our study was to identify gaps in awareness and behavior that may contribute to the development of diabetic foot complications and to propose strategies for improving patient care management and related outcomes.

## 2. METHODOLOGY and MATERIALS

### 2.1 Study Area

The study was carried out at the Medical-Outpatient Clinic(MOPC) in the St. Mary’s Mission Hospital, (StMMH), Nairobi, Kenya. The StMMH is a level 4 hospital and faith-based institution with a bed capacity of 320 and offers a variety of inpatient and outpatient general and specialty services, including emergency services, surgical, obstetrics & gynecology, pediatrics, gastroenterology, critical care, dental and maxillofacial, orthopedics, radiology and imaging, plus rehabilitation, ophthalmology, diabetes, hypertension, nutrition and comprehensive care clinics and HIV Counselling and Testing services among many other services. The busy MOPC attend to people living with various types of conditions including Diabetes mellitus on designated days and staffed by relevant clinicians. It is also a teaching hospital for neighboring medical institutions.

### 2.2 Study Design, Sampling Method, and Respondents

This research was descriptive cross-sectional study, based at a health institution and involved respondents (n= 140) who were recruited by census method at the Medical Outpatient Clinic of StMMH over one month of the study period. Census method was applied in this study where all adult diabetes patients who attended the clinic were enrolled. Since the study used census method, all adult patients with diabetes who gave consent to take part in the study were enrolled.

### 2.3 Data Instruments, Collection Method and Procedure

A pretested, structured questionnaire with both closed and open-ended questions was used to collect data. The closed-ended questionnaire was used to collect quantitative data including socio-demographics, dietary intake patterns, alcohol consumption and smoking habits, physical activity of the respondents, whereas the open-ended questionnaire was used to assess respondents present knowledge regarding blood glucose control, diabetic complications, adherence to feet examination and care, choice and care of shoes. The researcher adapted the questionnaire from the WHO STEP-wise approach to noncommunicable disease risk factor surveillance [6] for the purpose of this research study.

### 2.4 Validity and Reliability of the Study Tools

The project researcher adapted the questionnaire from the WHO STEP-wise approach to non communicable disease risk factor surveillance [6] for the purpose of this study.

Thereafter the assessment tool was evaluated for content validity to ensure they were complete and relevant to experts in endocrinology and in particular diabetes management. The data tool was pretested for validity and reliability at the MOPC / Mbagathi County Hospital in Nairobi. The researcher used 5% of the desired sample size in pretesting the tool.

### 2.5 Ethical Consideration

Ethical clearance for the study was obtained from the Kenyatta National Hospital –University of Nairobi Ethical Review Committee (KNH-UoN ERC) (approval number **UP 25/01/2021)**. We obtained a study permit from the National Commission for Science, Technology and Innovation (NACOSTI) before commencement of data collection, (approval number **NACOSTI/P/21/9811**). We also obtained institutional permission in writing from the St. Mary’s Mission Hospital in Nairobi to conduct the research in the facility. Informed consent was obtained from each study participant after clearly explaining the aim and goals of the study. Confidentiality regarding the study participants and their personal data was upheld throughout the study and data secured in a password protected computer.

Due to the prevailing Corona Virus Pandemic, public health safety measures were put in place to protect both the study participants and the researcher/ data collection assistants such as maintaining social distancing, use of face masks, use of hand sanitizers before and after attending to each participant and cough etiquette.

### 2.6 Data Analyses

Completed questionnaires were checked, cleaned and coded before entry of data. Responses from the open-ended questionnaires were categorized for qualitative information and then analyzed as quantitative data. A computer software, Statistical Package for Social Sciences (SPSS V. 25) was used for data analysis. Frequencies and percentages were obtained for categorical variables and for continuous variables. For inferential statistics, the chi-square test of independence was used to determine significance between independent and dependent categorical variables and a P-value of ≤0.05 was considered statistically significant. Data presentation was done in tables and figures

## 3. RESULTS

### 3.1 Demographic Characteristics of the Respondents

A total of n=140 participants were recruited in this study, majority of whom were above 60 years of age 50.0% (n=70) with more males 52.1% (n=73). A high proportion of the respondents were married 67.9% (n=95) while majority were Christians 87.9% (n=123). Those who had attained a secondary level of education was 45.7% (n=64) and unemployed 57.9% (n=81). Economically respondents with monthly income between KSh (Kenyan Shillings) 20001-60000 were 60.0% (n=84). Majority of the respondents 55% (n=77) reported ownership of housing, while more than two thirds 83% (n=117) had access to piped water. Half of the respondents 52% (n=73) had no medical insurance while 81.4% (n=114) sourced their food from markets as shown on Table 1 below:

**Table 1:** Socio-Demographic Characteristics of Respondents.

| Characteristics | Frequency (n) | Percent (%) |
| --- | --- | --- |
| **Age** |  |  |
| ≤ 50 years | 31 | 22.1 |
| 51 - 60 years | 39 | 27.9 |
| >60 years | 70 | 50.0 |
| **Gender** |  |  |
| Male | 73 | 52.1 |
| Female | 67 | 47.9 |
| **Marital status** |  |  |
| Single | 2 | 1.4 |
| Married/ Cohabiting | 95 | 67.9 |
| Divorced/ Separated/ Widowed | 43 | 30.7 |
| **Religion** |  |  |
| Christian | 123 | 87.9 |
| Muslim | 17 | 12.1 |
| **Level of education** |  |  |
| No formal education | 35 | 25.0 |
| Primary | 16 | 11.4 |
| Secondary | 64 | 45.7 |
| Tertiary | 25 | 17.9 |
| **Occupation** |  |  |
| Self-employed | 39 | 27.9 |
| Government employee | 11 | 7.9 |
| Non-government employee | 9 | 6.4 |
| Unemployed | 81 | 57.9 |
| **Monthly income (Kshs)** |  |  |
| Less than 20000 | 29 | 20.7 |
| 20001-60000 | 84 | 60.0 |
| Above 60000 | 27 | 19.3 |
| **Housing Arrangement** |  |  |
| Owned | 77 | 55.0 |
| Not owned | 63 | 45.0 |
| **Piped Water Access** |  |  |
| Yes | 117 | 83.6 |
| No | 23 | 16.4 |
| **Medical Insurance** |  |  |
| Has cover | 67 | 47.9 |
| Does not have cover | 73 | 52.1 |
| **Food access** |  |  |
| Market | 114 | 81.4 |
| Garden | 9 | 6.4 |
| Market and Garden | 17 | 12.2 |

### 3.2 Respondents Knowledge Level on Diabetic Foot Care

Majority of the respondents, 66% (n=93) demonstrated a low level of knowledge on diabetic foot care, followed by moderate knowledge level 26% (n=36), and only 8% n=11) had high knowledge level as shown in the Figure 1 below. The specific indices used in scoring are also illustrated in the Table 2 below.

**Figure 1:**
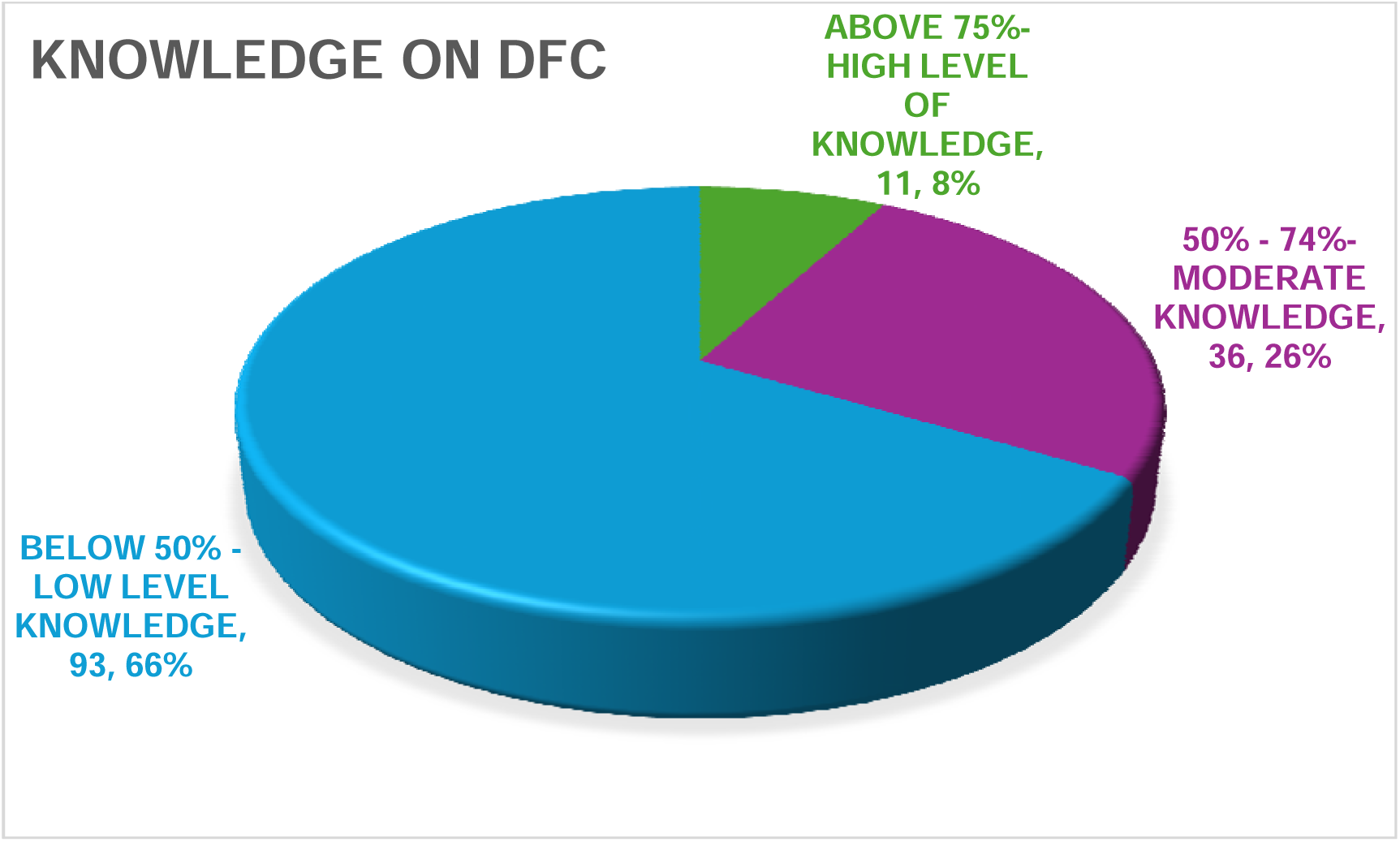
Respondents’ knowledge level on diabetic foot care

**Table 2:**
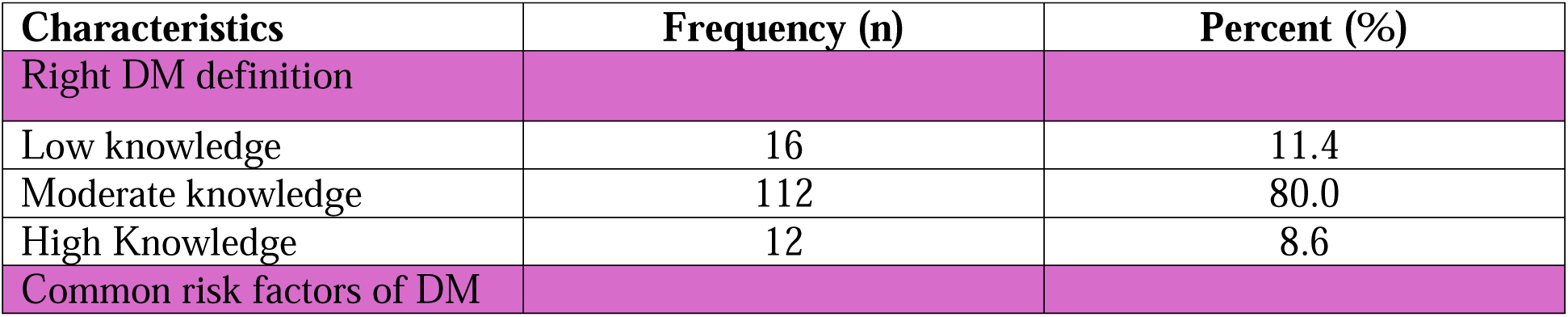
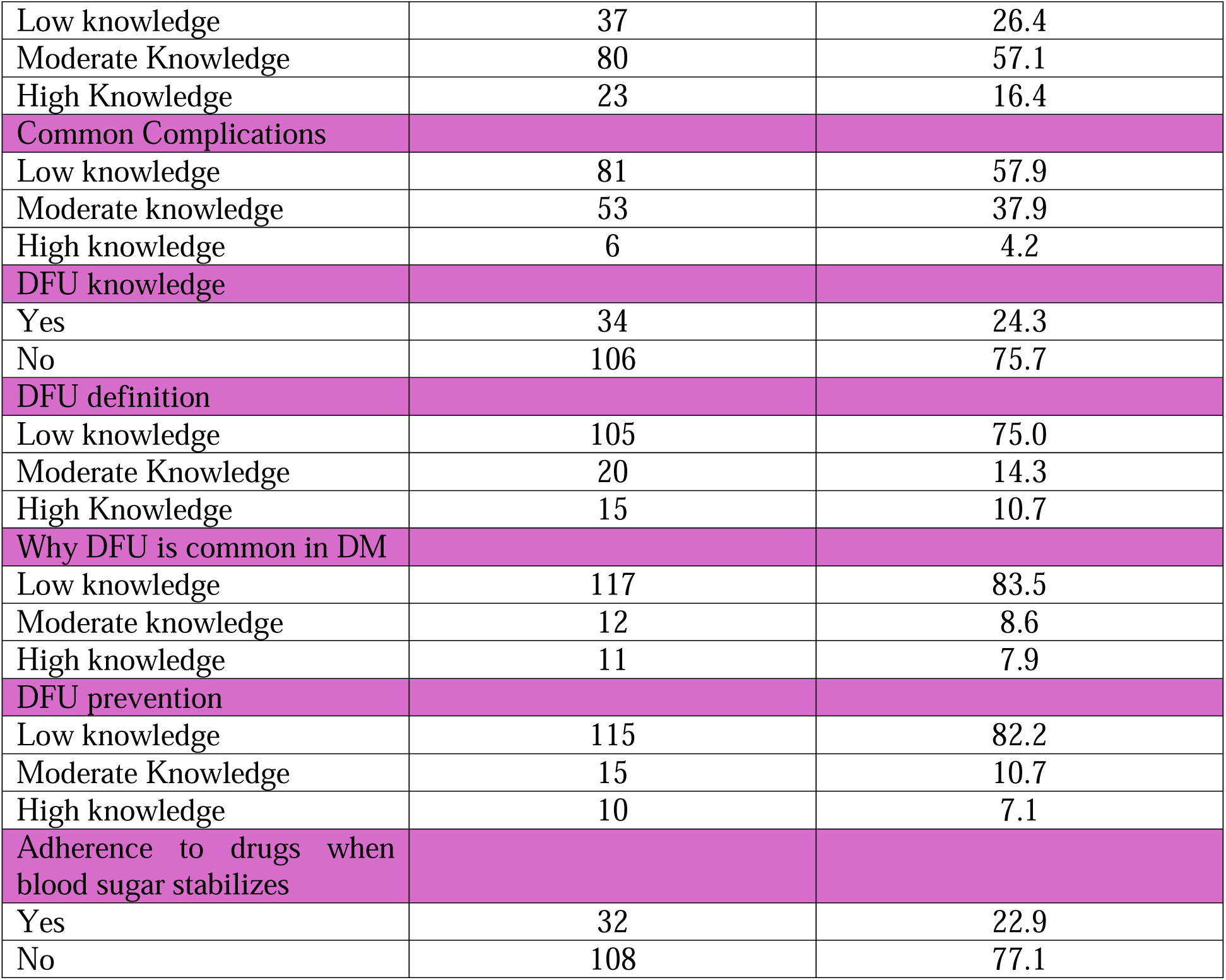
Respondents’ Knowledge of Diabetic Foot Care.

The Table 2 below shows respondents’ knowledge of specific areas of diabetes and diabetic foot care, where 8.6% (n=12) were able to correctly define diabetes mellitus, and 10.7% (n=15) had correctly defined diabetic foot ulcer. The majority reported no prior or first knowledge of diabetic foot ulcer 75.7% (n=106). The proportion of respondents who named risk factors associated with DM was 16.4% (n=23) and only 4.3% (n=6) could name the DM complications such as diabetes foot ulcers. A small proportion of the respondents 7.1% (n=10) reported high knowledge on DFU prevention while majority, 77.1% (n=108) reported good adherence to prescribed hypoglycemic medicines.

### 3.3 Respondents’ Practice Level on Diabetic Foot Care

The assessment of respondents’ practices included; dietary practices, social practices (smoking and alcohol consumption), diabetes drug adherence and foot care. The bar graph below demonstrates the practice level on diabetic foot care among the respondents (n, %) Figure 2.

**Figure 2:**
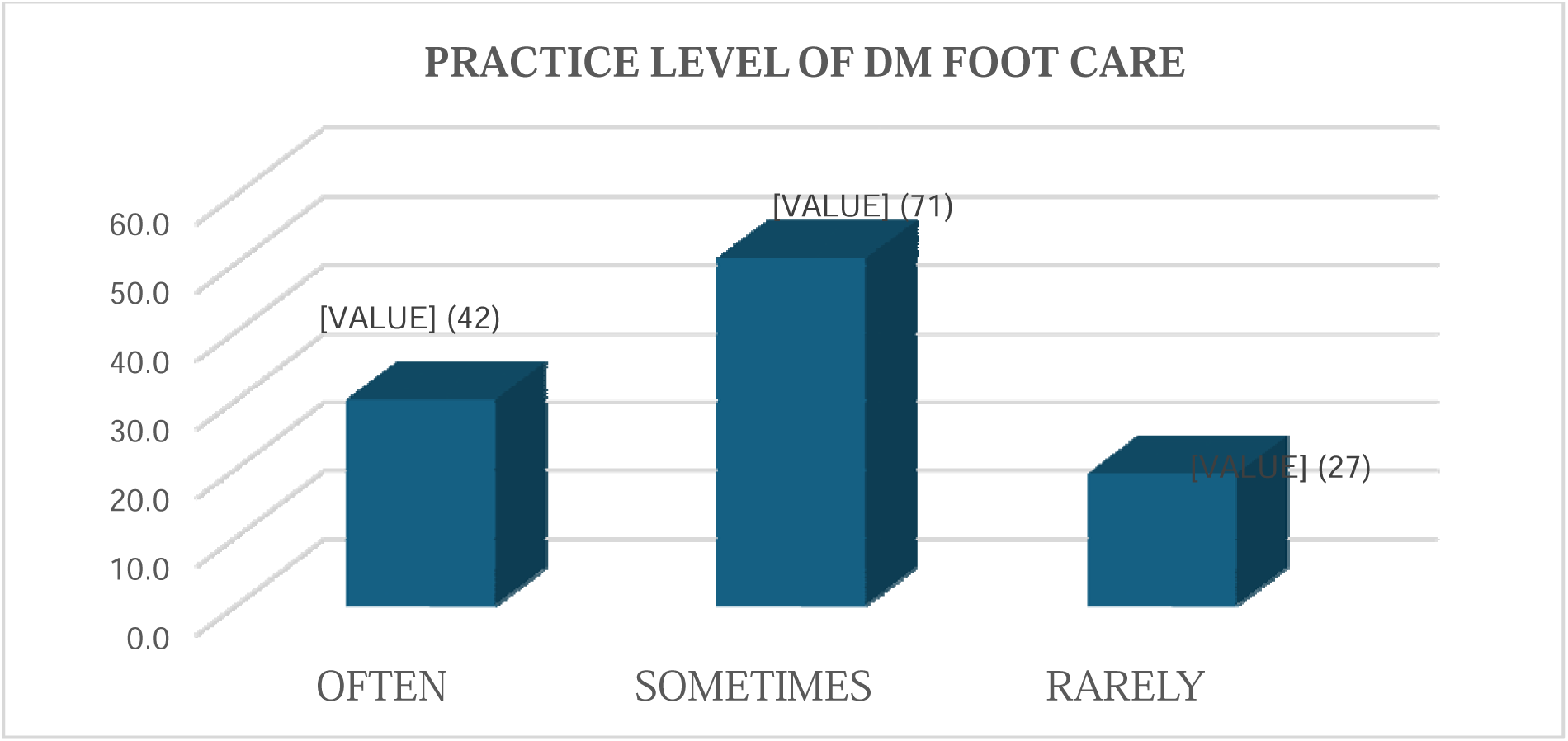
Respondents’ Practice Level on Diabetic Foot Care

#### 3.3.1 Respondents’ Dietary Practices Towards Diabetic Foot Care

Many of the respondents 84.3% (n=118) reported of consuming 3 meals a day, while only 2.8% (n=4) had more than five meals a day and the highest meal in quantity was dinner 52.9% (n=74) followed by lunch 32.1% (45). Majority of the respondents 66.4% (n=93), never take fast or processed foods and half of the respondents 53.6% (n=75) reported preference of whole meal/wheat bread to white bread. Majority of respondents 61.5% (n=86) consume legumes often/always, 72.9% (n=102) consume fruits 3-6 days a week and 62.2% (n=87) consumed vegetables 3-6 days a week. Majority of the respondents 83.6% (n=117), do not consume sugary snacks while 5% (n=7) of them consume sugary snacks often. Majority of them avoid sugar 87.9% (n=123) and 81.4% (n=114) of them do not add sugar to their beverages. Concerning salt consumption, majority of the respondents, 84.3% (n=118) reported use of salt in their diet, another 40% (n=56) use low sodium salt and 44% (n=31.4) reported using spices in meals. Majority of the respondents, 84.3% (n=118) use vegetable oil in cooking while few of them, 14.3% (n=20) did not know the type of oil used in cooking their meals (See Table 3).

**Table 3:** Dietary Practices of the Respondents.

| Characteristics | Frequency (n) | Percent (%) |
| --- | --- | --- |
| **Number of meals /days** |  |  |
| ≤3 Meals | 118 | 84.3 |
| 4 | 18 | 12.9 |
| ≥5 Meals | 4 | 2.8 |
| **Highest meal in frequency** |  |  |
| Breakfast | 21 | 15.0 |
| Lunch | 45 | 32.1 |
| Dinner | 74 | 52.9 |
| **Fast foods consumption** |  |  |
| Often | 9 | 6.4 |
| Rarely | 38 | 27.2 |
| Never | 93 | 66.4 |
| **Preferred bread** |  |  |
| Whole meal/brown | 75 | 53.6 |
| White | 6 | 4.3 |
| Both | 48 | 34.2 |
| None | 11 | 7.9 |
| **Legume consumption** |  |  |
| Always | 86 | 61.5 |
| Rarely | 45 | 32.1 |
| Never | 9 | 6.4 |
| **Fruit consumption** |  |  |
| Daily | 24 | 17.1 |
| 3-6 Days | 102 | 72.9 |
| < 3 Days | 14 | 10.0 |
| **Vegetable consumption** |  |  |
| Daily | 38 | 27.1 |
| 3-6days | 87 | 62.2 |
| < 3 days | 15 | 10.7 |
| **Type of cooking oil used** |  |  |
| Vegetable oil | 118 | 84.3 |
| Fats | 1 | 10.7 |
| None | 1 | 10.7 |
| Don't know | 20 | 14.3 |
| **Intake of sugary snacks** |  |  |
| Often | 7 | 5.0 |
| Sometimes | 16 | 11.4 |
| Never | 117 | 83.6 |
| **Read labels for sugar content** |  |  |
| Yes | 47 | 33.6 |
| No | 93 | 66.4 |
| Avoid sugary beverages |  |  |
| Yes | 123 | 87.9 |
| No | 17 | 12.1 |
| Sugar intake/cup of beverage |  |  |
| None | 114 | 81.4 |
| 1 Teaspoon | 24 | 17.2 |
| >1 Teaspoon | 2 | 1.4 |
| Salt use |  |  |
| Yes | 118 | 84.3 |
| No | 22 | 15.7 |
| Quantity of salt used |  |  |
| Pinch | 33 | 23.6 |
| Above teaspoon | 91 | 65.0 |
| Not aware | 16 | 11.4 |
| Limit processed foods |  |  |
| Yes | 117 | 83.6 |
| No | 23 | 16.4 |
| Buy low sodium salt |  |  |
| Yes | 56 | 40.0 |
| No | 84 | 60.0 |
| Use of spices |  |  |
| Yes | 44 | 31.4 |
| No | 96 | 68.6 |

#### 3.3.2 Social Practices Among Respondents

With regards to social practices and habits, most of the respondents 85.7% (n=120) reported of engaging in some form of exercise with most of them 68.6% (n=96) engaging in moderate physical activity and 13.6% (n=19) living a sedentary lifestyle. Majority of the respondents 64.3% n=90) exercise 3-4 days a week and half, 51.4% (n=72) exercise for less than 30 minutes a day. Concerning smoking habits, only 25% (n=35) had past smoking history and stopped, while 4.3% (n=6) were current smokers. Of the respondents, about a third, 30.7% (n=43) had a history of alcohol consumption while only 6.4% (n=9) were current alcohol consumers. Only 2.1% (n=3) of respondents consumed recommended standard rinks in a day, and majority did not respond to this question since they were not engaged in alcohol consumption 93.6% (n=131) as shown in Table 4 below:

**Table 4:** Respondents’ Lifestyle Practice of Foot Care.

| Characteristic | Frequency (n) | Percent (%) |
| --- | --- | --- |
| **Physical activity** |  |  |
| Yes | 120 | 85.7 |
| No | 20 | 14.3 |
| **Level of physical activity** |  |  |
| Vigorous | 18 | 12.9 |
| Moderate | 96 | 68.6 |
| Light | 7 | 5.0 |
| Sedentary | 96 | 13.6 |
| **Frequency of exercise** |  |  |
| ≥5 Days | 11 | 7.9 |
| 3-4 Days | 90 | 64.3 |
| 1-2 Days | 18 | 12.9 |
| None | 21 | 15.0 |
| **Duration per day** |  |  |
| ≥50 Minutes | 5 | 3.6 |
| 30-50 Minutes | 63 | 45.0 |
| ≤30 Minutes | 72 | 51.4 |
| **Smoking history** |  |  |
| Yes | 35 | 25.0 |
| No | 105 | 75.0 |
| **Currently smoking** |  |  |
| Yes | 6 | 4.3 |
| No | 134 | 95.7 |
| **Duration smoked** |  |  |
| ≥5 Years | 4 | 2.9 |
| <5 Years | 31 | 22.1 |
| N/A | 105 | 75.0 |
| **Alcohol use history** |  |  |
| Yes | 43 | 30.7 |
| No | 97 | 69.3 |
| **Currently drinking** |  |  |
| Yes | 9 | 6.4 |
| No | 131 | 93.6 |
| **Duration of alcohol use** |  |  |
| ≥5 years | 33 | 23.6 |
| <5 Years | 7 | 5.0 |
| N/A | 100 | 71.4 |
| Daily Standard drinks |  |  |
| ≤2 Bottles | 3 | 2.1 |
| ≥3 Bottles | 6 | 4.3 |
| N/A | 131 | 93.6 |

#### 3.3.3 Respondents’ Practice of Foot Care

Majority of the respondents 70% (n=98) do not soak their feet in warm soapy water while 90.7% (n=127) dry there intertoe spaces. Only 42.1% (n=59) use talcum powder and majority, 81.4% (n=114) had both trimmed nails and used moisturizers on feet. Majority 87.9% (n=123) use socks and shoes, and most of them 90.0% (n=126) inspect the inside of their shoes before wearing them as shown in table 5 below:

**Table 5:** Respondents’ Practice of Foot Care.

| Characteristics | Frequency (n) | Percent (%) |
| --- | --- | --- |
| **Soak in soapy water** |  |  |
| Yes | 42 | 30.0 |
| No | 98 | 70.0 |
| **Dry intertoe spaces** |  |  |
| Yes | 127 | 90.7 |
| No | 13 | 9.3 |
| **Use talcum powder** |  |  |
| Yes | 59 | 42.1 |
| No | 81 | 57.9 |
| **Trimmed nails** |  |  |
| Yes | 114 | 81.4 |
| No | 26 | 18.6 |
| **Use of moisturiser** |  |  |
| Yes | 114 | 81.4 |
| No | 26 | 18.6 |
| **Use of socks and shoes** |  |  |
| Yes | 123 | 87.9 |
| No | 17 | 12.1 |
| **Inspection of inner shoes before wearing** |  |  |
| Yes | 126 | 90.0 |
| No | 14 | 10.0 |

#### 3.3.4 Clinical Characteristics of Respondents

Regarding blood glucose measurements, 15 of the respondents had a fasting blood sugar reading and majority of them, 10.0% (n=14) had hyperglycemia, while majority, 54.3% (n=76) of those who did random blood sugar had hyperglycemia. Majority of the respondents 98.6% (n=138) had abnormal Waist-Hip Ratio (WHR) while half of respondents, 52.1% (n=73) were overweight as shown in Table 6 below:

**Table 6:** Clinical Characteristics of Respondents.

| Variable | Frequency(n) | Percent (%) |
| --- | --- | --- |
| **Fasting blood glucose** |  |  |
| Normal 4.0-9.0 | 1 | 0.7 |
| High>7.0 | 14 | 10.0 |
| **Random blood glucose** |  |  |
| Normal ≤11.1 | 49 | 35.0 |
| High >11.1 | 76 | 54.3 |
| **Blood pressure** |  |  |
| Normal | 55 | 39.3 |
| Pre-hypertensive | 31 | 22.1 |
| Hypertensive | 54 | 38.6 |
| **WHR** |  |  |
| Normal -Low risk | 2 | 1.4 |
| Abnormal-High risk | 138 | 98.6 |
| **BMI** |  |  |
| Underweight | 1 | 0.7 |
| Normal weight | 37 | 26.4 |
| Overweight | 73 | 52.1 |
| Obese | 29 | 20.8 |

### 3.4 Socio-Demographic Variables Associated with Diabetic Foot Care Among Adult Diabetic Patients at St. Mary’s Mission Hospital, Nairobi (n, %)

Using Chi square test, the proportion of failing to practice diabetic foot care on daily basis was significantly higher (X^2^ =7.514^a^, p = 0.023) among respondents who were married as compared to the rest. No significant association was found between the practice of diabetic foot care and age gender, religion, level of education, occupation and monthly income of the respondents. This is shown in Table 7 below:

**Table 7:** Relationship Between Socio-Demographic Variables and Frequency of Diabetic Foot Care Practices Among Respondents.

| Characteristics | Frequency of foot care |  |  | Chi(X<sup>2</sup>) | Df | p-value |
| --- | --- | --- | --- | --- | --- | --- |
|  | Daily | Not daily | Total |  |  |  |
| Age | n (%) | n (%) | n (%) |  |  |  |
| < 50 Years | 2(25) | 6(75) | 8(100) | 4.204<sup>a</sup> | 3 | 0.240 |
| 51 - 60 years | 6(26.1) | 17(73.9) | 23(100) |  |  |  |
| >61 years | 32(29.4) | 77(70.6) | 109(100) |  |  |  |
| Gender |  |  |  |  |  |  |
| Male | 21(28.8) | 52(71.2) | 73(100) | .003<sup>a</sup> | 1 | 0.957 |
| Female | 19(28.4) | 48(71.6) | 67(100) |  |  |  |
| Marital status |  |  |  |  |  |  |
| Single | 2(100.0) | 0(0.0) | 2(100) | 7.514<sup>a</sup> | 2 | 0.023 |
| Married | 30(31.6) | 65(68.4) | 95(100) |  |  |  |
| Divorced/ Separated/<br>Widowed | 8(18.6) | 35(81.4) | 43(100) |  |  |  |
| Religion |  |  |  |  |  |  |
| Christian | 35(28.5) | 88(71.5) | 123(100) | 0.007<sup>a</sup> | 1 | 0.935 |
| Muslim | 5(29.4) | 12(70.6) | 17(100) |  |  |  |
| Level of education |  |  |  |  |  |  |
| No formal education | 6(17.1) | 29(82.9) | 35(100) | 4.046<sup>a</sup> | 3 | 0.257 |
| Primary | 4(25.0) | 12(75.0) | 16(100) |  |  |  |
| Secondary | 23(35.9) | 41(100) | 64(100) |  |  |  |
| Tertiary | 7(28.0) | 18(72.0) | 25(100) |  |  |  |
| Occupation |  |  |  |  |  |  |
| Self-employed | 11(28.2) | 28(71.8) | 39(100) | 7.768<sup>a</sup> | 3 | 0.051 |
| Government employee | 7(63.6) | 4(36.4) | 11(100) |  |  |  |
| Non-government employee | 3(33.3) | 6(66.7) | 9(100) |  |  |  |
| Unemployed | 19(37.0) | 62(76.5) | 81(100) |  |  |  |
| Monthly income |  |  |  |  |  |  |
| ≤20000 | 7(24.1) | 22(75.9) | 29(100) | 5.729<sup>a</sup> | 3 | 0.126 |
| 20,001-60,000 | 23(27.4) | 61(43.6) | 84(100) |  |  |  |
| ≥ 61,000 | 10(37.0) | 17(63.0) | 27(100) |  |  |  |

### Demographic Characteristics of Respondents

Using Chi square test, there was no significant association between the knowledge on diabetic foot care and socio-demographic characteristics of respondents as shown on Table 8 below.

**Table 8:** Relationship Between Knowledge on Preventive Foot Care and Socio-Demographic Characteristics of Respondents.

| Characteristics | Knowledge level score | | | Total | Chi ($\chi^2$) | df | p-value |
| --- | --- | --- | --- | --- | --- | --- | --- |
|  | >75<br>high<br>level | 50-75<br>moderate | <50 low<br>level |  |  |  |  |
| **Age in years** | **n (%)** | **n (%)** | **n (%)** | **n (%)** |  |  |  |
| <50 | 0(0.0) | 11(35.5) | 17(54.8) | 31(100) | 7.403<sup>a</sup> | 6 | 0.285 |
| 51-60 | 5(12.8) | 8(20.5) | 26(66.7) | 39(100) |  |  |  |
| ≥60 | 3(4.3) | 17(24.3) | 50(71.4) | 70(100) |  |  |  |
| **Gender** |  |  |  |  |  |  |  |
| Male | 8(11.0) | 23(31.5) | 42(57.5) | 73(100) | 5.675<sup>a</sup> | 2 | 0.059 |
| female | 3(4.5) | 13(19.4) | 51(76.1) | 67(100) |  |  |  |
| **Marital status** |  |  |  |  |  |  |  |
| Single | 0(0.0) | 0(0.0) | 2(100.0) | 2(100) | 1.436<sup>a</sup> | 4 | 0.838 |
| Married/cohabiting | 8(8.40) | 26(27.4) | 61(64.2) | 95(100) |  |  |  |
| Divorced/separated/<br>widowed | 3(7) | 10(23.3) | 30(69.8) | 43(100) |  |  |  |
| **Religion** |  |  |  |  |  |  |  |
| Christian | 11(8.9) | 34(27.6) | 78(63.4) | 123(100) | 4.370<sup>a</sup> | 2 | 0.112 |
| Muslim | 0(0.0) | 2(11.8) | 15(88.2) | 17(100) |  |  |  |
| **Level of education** |  |  |  |  |  |  |  |
| No formal<br>education | 2(5.7) | 7(20.0) | 26(74.3) | 35(100) | 18.512<sup>a</sup> | 6 | 0.005 |
| Primary education | 2(5.7) | 7(20.0) | 26(74.3) | 35(100) |  |  |  |
| Secondary<br>Education | 2(5.7) | 7(20.0) | 26(74.3) | 35(100) |  |  |  |
| Tertiary education | 3(12.0) | 12(48.0) | 10(40.0) | 25(100) |  |  |  |
| **Monthly income** |  |  |  |  |  |  |  |
| ≤ 20,000 | 2(6.9) | 7(24.1) | 20(69.0) | 29(100) | 11.234<sup>a</sup> | 6 | 0.081 |
| 20,001-60,000 | 4(4.4) | 19(22.6) | 61(72.6) | 84(100) |  |  |  |
| ≥ 60,001 | 5(18.5) | 10(37.0) | 12(44.4) | 27(100) |  |  |  |

#### 3.4.2 Relationship Between Dietary Practices, Blood Sugar Levels, Physical Activity and Observed Foot Status Among Respondents

Using Chi square test, the prevalence of having intact feet was significant (X^2^ =12.227^a^, p = 0.000) among respondents who consume vegetables. The proportion of those with intact feet was significantly (X^2^ =5.499^a^, p = 0.0019), (X^2^ =5.146^a^, p = 0.023) higher among the respondents who are not currently taking alcohol and those who are not smoking currently respectively. No significant association was found between the observed foot status and fasting BS, Random BS, frequency of fruits intake, salt intake, sugar intake, physical activity and waist hip ratio. This is shown in Table 9 below:

**Table 9:**
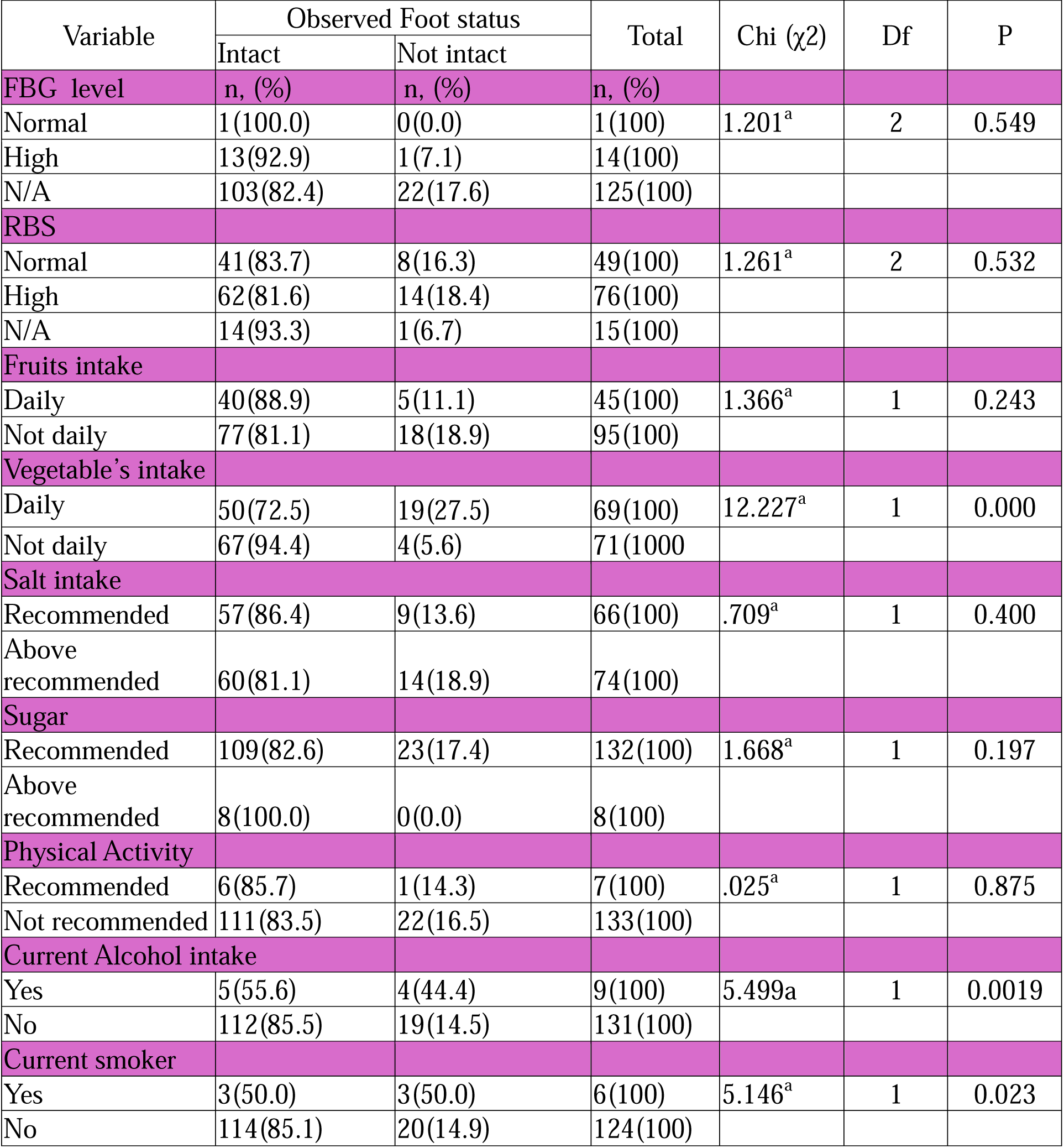
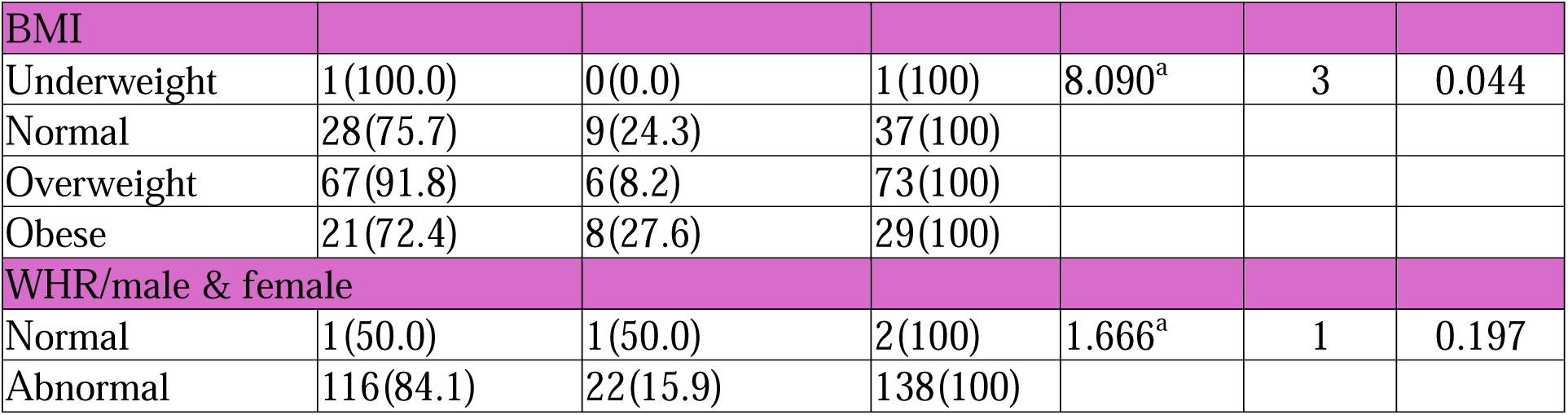
Relationship Between Dietary Practices, Blood Sugar Levels, Physical Activity and Observed foot Status Among Respondents.

| Variable | Observed Foot status | | Total | Chi ($\chi^2$) | Df | P |
| --- | --- | --- | --- | --- | --- | --- |
|  | Intact | Not intact |  |  |  |  |
| **FBG level** | n, (%) | n, (%) | n, (%) |  |  |  |
| Normal | 1(100.0) | 0(0.0) | 1(100) | 1.201<sup>a</sup> | 2 | 0.549 |
| High | 13(92.9) | 1(7.1) | 14(100) |  |  |  |
| N/A | 103(82.4) | 22(17.6) | 125(100) |  |  |  |
| **RBS** |  |  |  |  |  |  |
| Normal | 41(83.7) | 8(16.3) | 49(100) | 1.261<sup>a</sup> | 2 | 0.532 |
| High | 62(81.6) | 14(18.4) | 76(100) |  |  |  |
| N/A | 14(93.3) | 1(6.7) | 15(100) |  |  |  |
| **Fruits intake** |  |  |  |  |  |  |
| Daily | 40(88.9) | 5(11.1) | 45(100) | 1.366<sup>a</sup> | 1 | 0.243 |
| Not daily | 77(81.1) | 18(18.9) | 95(100) |  |  |  |
| **Vegetable's intake** |  |  |  |  |  |  |
| Daily | 50(72.5) | 19(27.5) | 69(100) | 12.227<sup>a</sup> | 1 | 0.000 |
| Not daily | 67(94.4) | 4(5.6) | 71(1000) |  |  |  |
| **Salt intake** |  |  |  |  |  |  |
| Recommended | 57(86.4) | 9(13.6) | 66(100) | .709<sup>a</sup> | 1 | 0.400 |
| Above recommended | 60(81.1) | 14(18.9) | 74(100) |  |  |  |
| **Sugar** |  |  |  |  |  |  |
| Recommended | 109(82.6) | 23(17.4) | 132(100) | 1.668<sup>a</sup> | 1 | 0.197 |
| Above recommended | 8(100.0) | 0(0.0) | 8(100) |  |  |  |
| **Physical Activity** |  |  |  |  |  |  |
| Recommended | 6(85.7) | 1(14.3) | 7(100) | .025<sup>a</sup> | 1 | 0.875 |
| Not recommended | 111(83.5) | 22(16.5) | 133(100) |  |  |  |
| **Current Alcohol intake** |  |  |  |  |  |  |
| Yes | 5(55.6) | 4(44.4) | 9(100) | 5.499<sup>a</sup> | 1 | 0.0019 |
| No | 112(85.5) | 19(14.5) | 131(100) |  |  |  |
| **Current smoker** |  |  |  |  |  |  |
| Yes | 3(50.0) | 3(50.0) | 6(100) | 5.146<sup>a</sup> | 1 | 0.023 |
| No | 114(85.1) | 20(14.9) | 124(100) |  |  |  |
| **BMI** |  |  |  |  |  |  |
| Underweight | 1(100.0) | 0(0.0) | 1(100) | 8.090<sup>a</sup> | 3 | 0.044 |
| Normal | 28(75.7) | 9(24.3) | 37(100) |  |  |  |
| Overweight | 67(91.8) | 6(8.2) | 73(100) |  |  |  |
| Obese | 21(72.4) | 8(27.6) | 29(100) |  |  |  |
| **WHR/male & female** |  |  |  |  |  |  |
| Normal | 1(50.0) | 1(50.0) | 2(100) | 1.666<sup>a</sup> | 1 | 0.197 |
| Abnormal | 116(84.1) | 22(15.9) | 138(100) |  |  |  |

#### 3.4.3 Frequency of Fruits and Vegetables Intake in Relation to Blood Sugar (N, %)

Using Chi Square test of significance, no significant relationship was found between blood sugar levels and the frequency of fruits and vegetable intake among the respondents as shown in the Table 10 below:

**Table 10:**
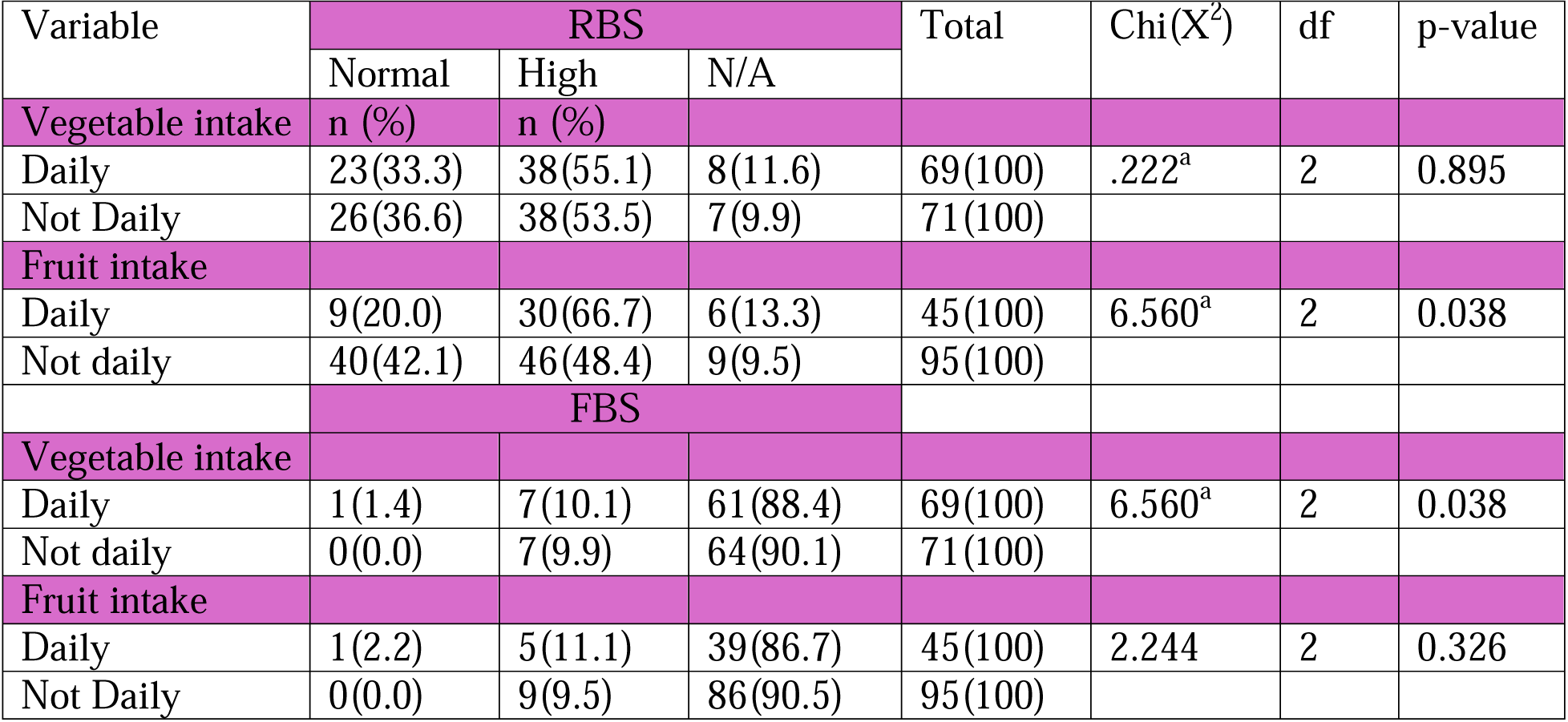
Relationship Between Frequency of Fruits and Vegetables Consumption and Blood Sugar.

| Variable | RBS |  |  | Total | Chi(X<sup>2</sup>) | df | p-value |
| --- | --- | --- | --- | --- | --- | --- | --- |
|  | Normal | High | N/A |  |  |  |  |
| **Vegetable intake** | **n (%)** | **n (%)** |  |  |  |  |  |
| Daily | 23(33.3) | 38(55.1) | 8(11.6) | 69(100) | .222<sup>a</sup> | 2 | 0.895 |
| Not Daily | 26(36.6) | 38(53.5) | 7(9.9) | 71(100) |  |  |  |
| **Fruit intake** |  |  |  |  |  |  |  |
| Daily | 9(20.0) | 30(66.7) | 6(13.3) | 45(100) | 6.560<sup>a</sup> | 2 | 0.038 |
| Not daily | 40(42.1) | 46(48.4) | 9(9.5) | 95(100) |  |  |  |
|  | **FBS** |  |  |  |  |  |  |
| **Vegetable intake** |  |  |  |  |  |  |  |
| Daily | 1(1.4) | 7(10.1) | 61(88.4) | 69(100) | 6.560<sup>a</sup> | 2 | 0.038 |
| Not daily | 0(0.0) | 7(9.9) | 64(90.1) | 71(100) |  |  |  |
| **Fruit intake** |  |  |  |  |  |  |  |
| Daily | 1(2.2) | 5(11.1) | 39(86.7) | 45(100) | 2.244 | 2 | 0.326 |
| Not Daily | 0(0.0) | 9(9.5) | 86(90.5) | 95(100) |  |  |  |

## DISCUSSION

In this study, we had more males than female participants. Half of the respondents were aged 60 years and above and a higher proportion of the participants were married, while majority were Christians and unemployed. This is contrary to a study conducted in Ethiopia where majority of the respondents (55.9%) were male contrary to religion where 78.9% were Muslims and 44.1% were self-employed as farmers [24]. This could be due to the differences in geography and socio-cultural contexts in the study population with a different study set up. There was no significant association found between the practice of diabetic foot care and age, gender, religion, level of education, occupation and monthly income among the respondents

According to this study, a majority of the participant, had abnormal waist hip ratio while half of respondents were overweight. This is closely related to a study in Iran where only few had normal body mass index while some had normal waist hip ratio [3] and this difference could be due to differences in dietary practices in the study population compared to the Persian diet. Also, half of the participants in this study, were above 60 years and ate their heaviest meal at dinner. Studies have shown that eating heavy meals last thing before bedtime is associated with risk factors of increased body mass index revealed by overweight/obesity and increased waist hip ratio among others. This can be complicated by slowed metabolism associated with increase in age. These risks factors increase the likelihood of developing foot problems especially among people living with diabetes.

Studies have pointed out that appropriate and effective self-foot care practices are an important aspect of preventing diabetes foot ulcers that every person living with diabetes should do continuously. According to our study, majority of the respondents do not soak their feet in warm soapy water, while majority dry their inter toe spaces after feet hygiene. Only less than half use talcum powder, and majority had both trimmed nails and used moisturizers on their feet.

Majority of the participants use socks and shoes and inspect the inside of their shoes before wearing them These findings agree with a study carried out in Iran that reported poor foot care practices among patients with diabetes specifically, majority do not walk barefoot and few use talcum powder or other powders between the toes, few had properly trimmed toenails and had kept their feet skin soft and intact [3]. Another study reported poor practices toward regular inspection of feet among patients in Qatar [25]. It also agrees with another study from Malaysia where few patients newly diagnosed with diabetes practiced good habits towards foot care [26]. In another study by Desalu and cohort from Nigeria, observed that only few of the patients with diabetes had good foot care practices [27].

Generally, this study found out that the practice of foot care among the participants studied was unsatisfactory. This shows that there is lack of adequate health education in the hospitals. Thus, a call for health care providers particularly nurses and hospital managers to design strategies in order to raise patients’ awareness of foot care practices to prevent morbidity and mortality associated with diabetic foot ulcers.

The prevalence of failing to practice diabetic foot care on daily basis was significantly higher among respondents who were married, which is contrary to a study in Jimma Medical Center where there was no significant difference in the proportion between male and female, single or married respondents in practicing the standard foot-care practices [28]. This could be associated with the low level of knowledge among the study participants regarding diabetes and diabetes foot care Considering that the research study was done during the Covid-19 pandemic, it is possible that individual patients followed the social distancing guidelines that denied them the benefit of practicing daily and regularly the basic foot care activities with the help of their family members. Our study found out that, majority of the participants demonstrated a low level of knowledge on diabetic foot care while few participants had high knowledge. These findings are like a study conducted in Iran, Saudi Arabia and Tobago, indicating that most of the participants had a poor knowledge [3,29,30] respectively. This is probably because there was no structured health educational talks on diabetes and diabetic foot practices that were being carried out at the hospital facility mostly because of the Covid-19 pandemic guidelines especially social distancing. This could also be attributed to recall bias during a life-threatening pandemic scenario where patients could easily define diabetes but could not list associated factors or complications.

There was no significant association revealed by this study between the knowledge on diabetic foot care and socio-demographic characteristics associated with diabetes foot care. This finding is like a study done in Kermanshah [30], but disagrees with a study conducted in Iran that observed that gender, duration of disease, occupation, place of residence, level of education had significant relationships with knowledge [3]. This is probably because of differences in study population characteristics and difference in the study set up.

### Limitation of the Study

It would have been even better to do the biochemical measurements of glycemic control among these respondents to assess any association as the random and fasting blood sugars did not show any significant association. This study was carried out in an urban, low-income setting, and, therefore, should be used with good caution to generalize its findings for either rural setting or urban, high-end settings. By its nature, a cross-sectional study will not demonstrate the cause-effect relationship among the study variables.

## CONCLUSION

According to our study, the level of knowledge of potential harm from diabetic mellitus was found to be low among the participants. Furthermore, the level of diabetic foot care was unsatisfactory. Additionally, most of the respondents consumed their heaviest meal at dinner placing them at risk of increased body mass index (BMI) and consequent risk of diabetic foot ulcers (DFU), plus other complications. Our work revealed that both levels of knowledge and practice towards foot care was not satisfactory. Therefore, we highly recommend that healthcare providers, particularly the nurses, be involved in designing and providing appropriate health packages for patients with diabetes -- instilling provisions of knowledge and foot care practices.

## ABBREVIATIONS

DM: Diabetes Mellitus
DFC: Diabetic Foot Care
DFU: Diabetic Foot Ulcers
KsH: Kenyan Schilling (Kenyan Currency)
MOPC: Medical-Outpatient Clinic
NACOSTI: National Commission for Science, Technology and Innovation
StMMH: Saint Mary’s Mission Hospital in Nairobi, Kenya
WHR: Waist-Hip Ratio

## Data Availability

All data produced in the present work are contained in the manuscript

## ACKNOWLEDGEMENTS

We offer our special thanks to the many health professionals at the St. Mary’s Mission Hospital in Nairobi who untiringly contributed to our research project, and we offer our appreciation to the adults who participated in our project, without whom our study would not have materialized. Also, we thank the dedicated project assistants who handled data collection with timely data curation.

## AUTHOR CONTRIBUTIONS

**Norah Anne Mogute Oyagi**: Co-Conceptualization, Project Administration, Formal Analysis, Writing – Original Draft, and Draft Reviewing & Editing.

**Okubastion Tekeste Okube**: Co-Conceptualization, Formal Analysis, and Validation.

**Magdalene Philip Umoh**: Validation, Data Review and Draft Reviewing & Editing.

**Abraham Isiaka Jimmy**: Methodology, Data Review, and Draft Reviewing & Editing.

**Lee Presley Gary, Jr**.: Resources, Visualization, and Manuscript Review & Editing.

## TRANSPARENCY

The authors volunteer that no *ChatGPT* or any equivalent AI program was not a source of data or information and was not used to draft or embellish this manuscript.

## AVAILABILITY of DATA and MATERIALS

Data for this study are openly available from the Lead Author (Norah Anne Mogute Oyagi) or the Corresponding Author (Lee Presley Gary, Jr.) upon a written or electronic request from any interested correspondent.

All relevant, collected data are presented in this manuscript. In case additional data is needed, it can be availed by upon reasonable written or electronic request.

## ETHICS APPROVAL and CONSENT to PARTICIPATE

Full ethical clearance for the research study was obtained from the Kenyatta National Hospital – University of Nairobi / Ethical Review Committee with Approval Permit Number **UP 25/01/2021)**.

Also, the authors received a study permit from the National Commission for Science, Technology and Innovation (NACOSTI) before commencement of any data collection: Approval Permit Number **NACOSTI -- P/21/9811**.

We sought and were granted institutional permission from St. Mary’s Mission Hospital in Nairobi to conduct our research project on its premises and to use its facility as proper.

Informed consent was obtained from each study participant after clearly explaining the aim and goals of the study and their vital role in the project. Confidentiality for the study participants and their personal data was upheld throughout the study -- and data was secured in a password protected computer.

## BIOGRAPHIES

**Norah Anne Mogute Oyagi** Is currently the Lead Nurse/ Matron at Holy Spirit Hospital in Makeni, Sierra Leone. She is a healthcare management specialist with 25 years of experience in Nursing practice and nursing education, coupled with hospital management. She has served in both urban and rural settings in Kenya, Ghana and Sierra Leone. She has keen interest in nursing education, public health, research, health systems management, health economics and healthcare financing, health policy and quality management to impact access to health and wellness.

**Okubastion Tekeste Okube** is a Senior Nurse Lecturer at Amref International University (AMIU), located in Nairobi, Kenya. His background covers over 12 years of dedicated experience in teaching nursing, improving clinical settings, community health projects, and health research and policy. He specializes in building clinical and facility care teams that deliver dedicated service, focusing on active training and personal mentorship. Tekeste holds a Ph.D. degree in nursing and thrives on problem solving and organizational management.

Abraham Isiaka Jimmy is a Public Health Specialist and Re-searcher. He has more than six years of experience working in emergency and development contexts in Sierra Leone and has developed strong competencies in health program management. He teaches public health and mental health courses at the University of Makeni, located in the City of Makeni in Sierra Leone. He advises on the operationalization of One Health Programs, including developing policies, strategies, and governance for mitigating public health challenges.

**Magdalene Philip Umoh** is a Public Health Scholar with expertise in Occupational/Environmental Health and Safety. She is a faculty lecturer for the Department of Public Health at the University of Makeni, located in Makeni, Sierra Leone, and serves as Director of the Center of Excellence in Maternal and Child Health Education and Research.

Her vast experience with public and community health issues covers over ten years. including lecturing, researching, and consulting with the Food and Agricultural Organization (FAO), Ministry of Agriculture, and various Human Capital Development Plans (HCD Plus). She is passionate about maternal and child care issues..

**Lee Presley Gary, Jr.** is a Visiting Research Scholar at the University of Makeni, located in the City of Makeni in Sierra Leone, and he is Owner/CEO of Strategic Management Services – USA, a global consultancy specializing in mitigating public health threats and hazards associated with dirty water and human waste, and served as a Fulbright Specialist at the University of Makeni teaching public health courses during 2024. He is an active member of the Water Environment Federation and is an adjunct instructor for the National Disaster Emergency Management University (Emmitsburg, MD). He was named a Fulbright Scholar at the University of Malta for 2025-2026.

## RESEARCH FIELDS

**Norah Anne Mogute Oyagi**: Nursing Research, Healthcare Management, Nursing Care Processes, Professional Nursing Education and Capacity Building, Human Resources for Healthcare, and Non-Communicable Diseases with keen interest in research on metabolic syndrome disorders seeking better outcomes for patients.

**Okubastion Tekeste Okube: C**ardiovascular Diseases, Maternal and Child Healthcare, Midwifery Sciences, Nursing Education, Ethical Issues in Nursing, Community Health, and Public Health Issues.

**Magdalene Philip Umoh:** Maternal Healthcare, Occupational Health Safety & Regulations, Environmental Health Risk & Assessments, Public Health Policy, Infectious Diseases, and Nursing Education & Career Development.

**Abraham Isiaka Jimmy:** Public Health Policy, Health System Financing, Healthcare Economics, Mental Health Policy, Maternal and Child Healthcare, Occupational Health, and Environmental Health Risks & Assessments.

**Lee Presley Gary, Jr.:** Disaster Management, Emergency Management, Mitigation of Dirty Water and Human Waste (WASH), and Case Studies for Public Health Issues.

* * * * *

